# Cooperative virus propagation in COVID-19 transmission

**DOI:** 10.1101/2020.05.05.20092361

**Authors:** Ziwei Dai, Jason W. Locasale

## Abstract

The global pandemic due to the emergence of a novel coronavirus (COVID-19) is a threat to humanity. There remains an urgent need to understand its transmission characteristics and design effective interventions to mitigate its spread. In this study, we define a non-linear (known in biochemistry models as allosteric or cooperative) relationship between viral shedding, viral dose and COVID-19 infection propagation. We develop a mathematical model of the dynamics of COVID-19 to link quantitative features of viral shedding, human exposure and transmission in nine countries impacted by the ongoing COVID-19 pandemic, and state-wide transmission in the United States of America (USA). The model was then used to evaluate the efficacy of interventions against virus transmission. We found that cooperativity was important to capture country-specific transmission dynamics and leads to resistance to mitigating transmission in mild or moderate interventions. The behaviors of the model emphasize that strict interventions greatly limiting both virus shedding and human exposure are indispensable to achieving effective containment of COVID-19. Finally, in the USA we find that by the summer of 2021, a difference of about 1.5 million deaths may be observed depending on whether the interventions are to be maintained strictly or lifted in entirety.

## Introduction

A novel disease (COVID-19)^1-3^, which was first detected in December 2019 and then found to be caused by a newly discovered coronavirus, severe acute respiratory syndrome coronavirus-2 (SARS-CoV-2), has become a global pandemic, causing millions of infections, and claiming more than 240,000 lives as of the time of this publication. In response to the outbreak of COVID-19, some countries such as China launched extremely aggressive, unprecedented actions including social distancing measures, school and workplace closures, travel bans, centralized quarantine of infected individuals, and contact tracing using cell phone tracking. While new infections and deaths have been greatly reduced in China^4-6^ and in some other countries, the disease is rapidly spreading in many countries, among which the United States, with mixed efforts to limit transmission, has suffered from the highest number of confirmed cases and deaths. It speaks for itself the importance and urgency of understanding what interventions may best serve public health. A knowledge about the underlying mechanisms governing virus transmission dynamics gives insight into this question.

Mathematical models have been used to understand the transmission dynamics of epidemics^7^ from seasonal influenza^8^ to the Ebola virus^9^, forecast their outbreak and spread, and evaluate the efficiency of interventions in containing these diseases^10^. The most widely applied mathematical models in epidemiology are compartmental models, for instance, the susceptible-infected-recovered-deceased (SIRD) model, in which the total population is divided into several categories susceptible, infected, recovered, and deceased, between which exchange occurs. These models have served a fundamental role in reaching a quantitative understanding of transmission characteristics of COVID-19, such as estimation of basic reproduction number (R_0_),incubation period, and fatality rate^11-15^, and evaluation of the efficacies of interventions^5,6,16-19^. While these models have greatly contributed to our current understanding of the COVID-19 pandemic, they lack information on infection mechanisms by SARS-CoV-2 and overlook the quantitative relationship between the rate of infection and viral exposure such as what would happen during a super-spreader event. Instead, spread of the disease is taken to be directly mediated by interactions between infected and susceptible individuals, which are assumed to occur through linear relationships, thus creating a gap between the modeling tools currently available and the knowledge about detailed parameters of the virus, such as rates of virus shedding and decay^20,21^, source-dependent viral dose^22,23^, and dose dependent infectivity^24^, for which experimental data have been collected but not yet used in the development of quantitative models of the pandemic^25^.

Here, we develop a novel mathematical model for the transmission dynamics of COVID-19. We show that the cooperativity in infection between uninfected and infected individuals is critical in explaining the transmission dynamics of COVID-19 in countries most severely impacted by the pandemic. Cooperativity has a long history in the study of biological dynamics that began with models of ligand binding to receptors with multiple binding sites and has numerous applications to understand fundamental molecular processes in living cells such as gene transcription and signal transduction^26,27^. Despite the complexity in the pathology of diseases such as COVID-19, there exists a mechanistic similarity between the binding of a ligand and activation of a receptor with multiple binding sites, and the release of virus from an infected person and transmission to an uninfected host which can be multiplicative through many avenues. Transmission can amplify from one person through airborne transmission and other dissemination of infectious liquid droplets. Preliminary studies on the dose-dependent response to virus infection have also demonstrated that the relationship between viral exposure and infection is non-linear, suggesting that the infection kinetics is more complicated than assumed in the original epidemic models^24^. Thus, consideration of such non-linearity may offer novel insights about how the virus is transmitted, how the pandemic can be effectively controlled, and how much the current public health interventions have contributed to its containment or lack thereof.

Our mathematical models indicate that, the transmission of COVID-19 is intrinsically resistant to weak-to-moderate interventions because of the cooperativity of virus infection, thereby requiring only aggressive interventions to mitigate its spread. As an example, evaluation of the COVID-19 transmission characteristics in the USA shows that the current schemes of interventions, which were temporarily strong but on average weak-to-moderate, may have saved over 500,000 lives as of July 15, 2020, and will have saved over 2.5 million lives by June 2021 if the interventions are to be maintained at their peak strength. The early lifting of interventions is unfortunately predicted to cost about 1.5 million lives and the COVID-19 pandemic in the United States is unlikely to be contained by current interventions without a large loss of human life.

## Results

### A cooperative infection model correctly captures country-specific COVID-19 transmission dynamics

To incorporate viral dose and cooperative infection into compartmental infectious disease models, we introduced a variable V quantifying the abundance of the virus to a reduced SIRD model in which the total number of recovered (R) and deceased (D) people are treated as a single variable, non-susceptible (N) population (Figure 1A, Methods). The reason for using a reduced model instead of the original SIRD model is that the rates for the transition from infected (I) to recovered (R) and deceased (D) are both proportional to the number of infected individuals (I), thus the sum of these two, N, also changes with a rate proportional to I. Moreover, a model with fewer parameters has the advantage of reducing parameter uncertainty and better avoiding overfitting. Using a standard mathematical term describing the cooperative relationship between V and infection of susceptible individuals by viable virus particles, we derived three variations of the original SIRD model, including the reduced SIRD model, the reduced SIRD model with virus pool, and the reduced SIRD model with virus pool and cooperative infection (Figure 1A, Methods). Cooperativity was modeled using a Hill function with two parameters accounting for the levels of cooperativity and saturation in response to viral dose, respectively (Methods). These models were then compared as to their accuracy in fitting data consisting of COVID-19 case numbers, recovered and deceased individuals in the nine countries with the highest and still rapidly increasing numbers of COVID-19 infections and deaths, which indicate ongoing COVID-19 pandemic as of Apr 3, 2020. These countries include USA, Turkey, UK, Spain, Italy, Germany, France, Switzerland, and Iran. Accuracy of fit was determined by a cost function defined as the mean squared deviation between simulated and actual daily numbers of infected individuals and non-susceptible individuals on dates within the time window between the confirmation of the first infection in the country and Apr 3, 2020 (Figure 1B). We found that among the three models, the reduced SIRD model with a virus pool and cooperative infection (i.e. the cooperative infection model) reached higher accuracy compared to the other two models in all countries, and was able to generate satisfactory fitting (i.e. optimal cost function < 0.005, which means that the mean error for each data point is less than 7.07% of the maximal value of actual data over the entire time course) in 7 out of the 9 countries (Figure 1C). By contrast, the two models without cooperative infection generated satisfactory fitting in only two countries, and the addition of the virus pool to the reduced SIRD model did not increase the fitting accuracy of the model despite the increased complexity of model (absolute difference in −log10(Optimal value of cost function) smaller than 0.003 for all countries, Figure 1C), suggesting that consideration of cooperativity in infection is critical for the model to accurately predict the COVID-19 transmission dynamics. We also performed a sensitivity analysis assuming that 50% or 80% of the infections were undocumented and repeated all analyses using the adjusted data, to confirm that the model is consistent with the COVID-19 transmission dynamics and that all conclusions are unaffected in the presence of reporting or testing biases (Methods). The cooperative infection model also includes parameters directly related to other factors contributing to the viral dose dynamics, such as rate constants for virus shedding and decay, and parameters quantifying the overall susceptibility of healthy population to the virus and efficiency of patient clearance (Figure 1D). Inference of these parameters from COVID-19 transmission data in different countries would offer valuable insights into the intrinsic properties of the virus and its specific transmission characteristics in each country.

**Figure 1.**
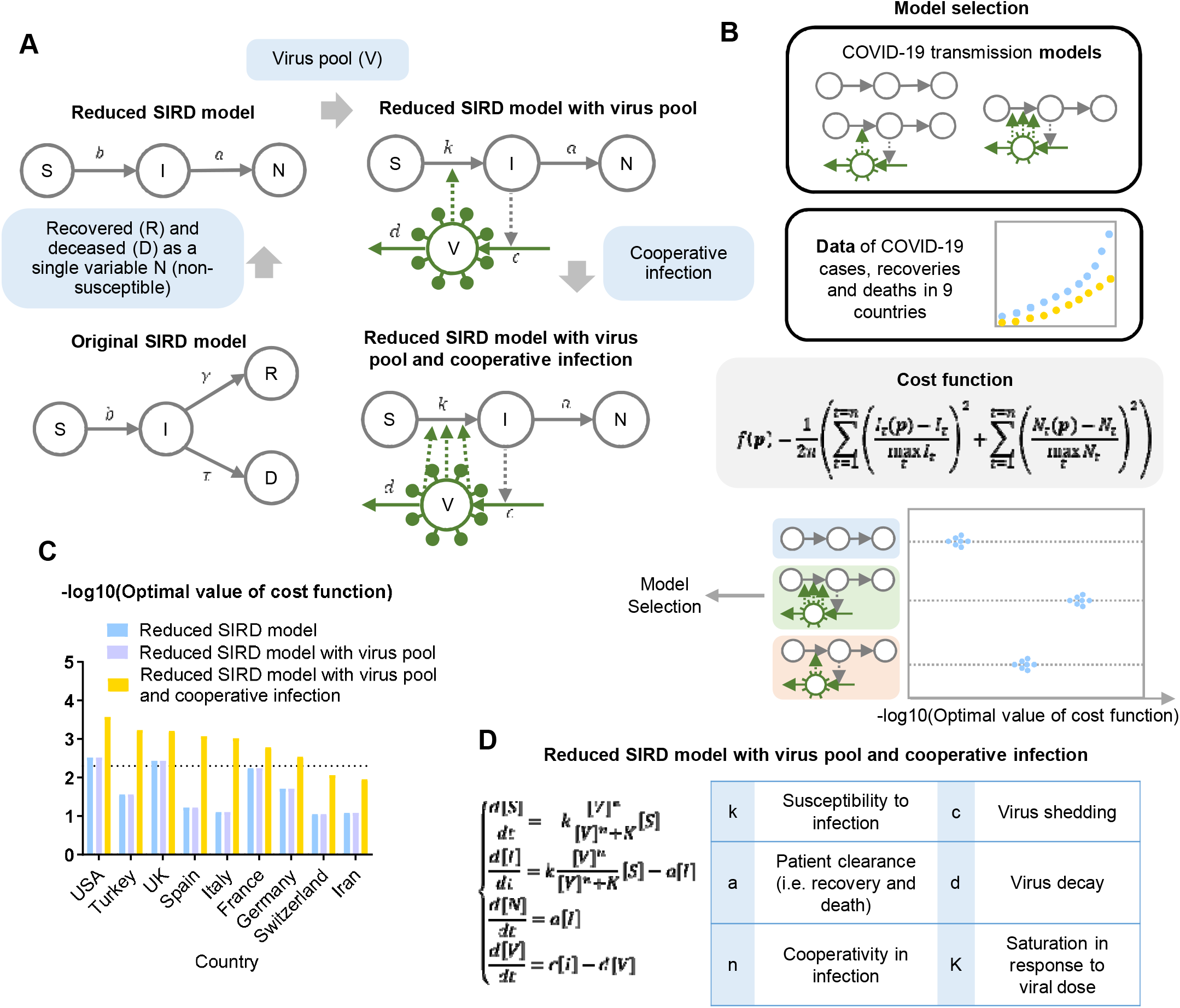
A cooperative infection model captures country-specific COVID-19 transmission dynamics. A. Diagrams of the original SIRD model, reduced SIRD model, reduced SIRD model with virus pool, and reduced SIRD model with virus pool and cooperative infection. B. Workflow for the model selection process and the definition of cost function used. C. Optimal cost function values achieved in fitting country-specific COVID-19 transmission data using the reduced SIRD model, reduced SIRD model with virus pool, and reduced SIRD model with virus pool and cooperative infection. The dashed horizontal line indicates the threshold of cost function value for satisfactory fitting. A parameter set with cost function value lower than this threshold was considered satisfactory. D. Equations for the reduced SIRD model with virus pool and cooperative infection and meaning of model parameters.

### The cooperative infection model predicts country-specific COVID-19 transmission dynamics

To predict COVID-19 transmission dynamics in the seven countries where the cooperative infection model was consistent with its previous transmission dynamics, we simulated the model over a time window of 80 days starting from February 15, 2020 with parameter sets that satisfactorily fit the data for each country. Parameter sets generating satisfactory fits were obtained by sampling the parameter space (Methods). We found that, in the absence of intervention, all seven countries were predicted to undergo significant increase in the number of infected individuals, especially for the countries UK, USA and Turkey, for which the numbers of infections were predicted to increase rapidly without any tendency of slowdown (Figure 2A, Supplementary Figure 1). To understand COVID-19 transmission characteristics in different countries, especially those predicted to face more future challenges, we examined distributions of parameters determining the transmission dynamics in each country (Figure 2B). We found that the two model parameters that significantly differ across countries are related to the susceptibility of the healthy population to infection (k) and the efficiency of patient clearance (a), while parameters quantifying viral dose dynamics (c and d) and the cooperativity in response to viral dose (n and K) showed no difference between countries (Figure 2B). These results further support the use of the cooperative infection model because parameters for virus shedding, decay and dose response are directly determined by intrinsic properties of the virus^20^, thus we expect values of these parameters to be indistinguishable between countries, while the susceptibility of the healthy population and efficiency of patient clearance reflects the performance of public health measures and the healthcare systems which are country-specific. Notably, USA, UK and Turkey are the three countries with lowest efficiency of patient clearance among the seven countries, while the USA population was also found to suffer from the highest susceptibility to infection.

**Figure 2.**
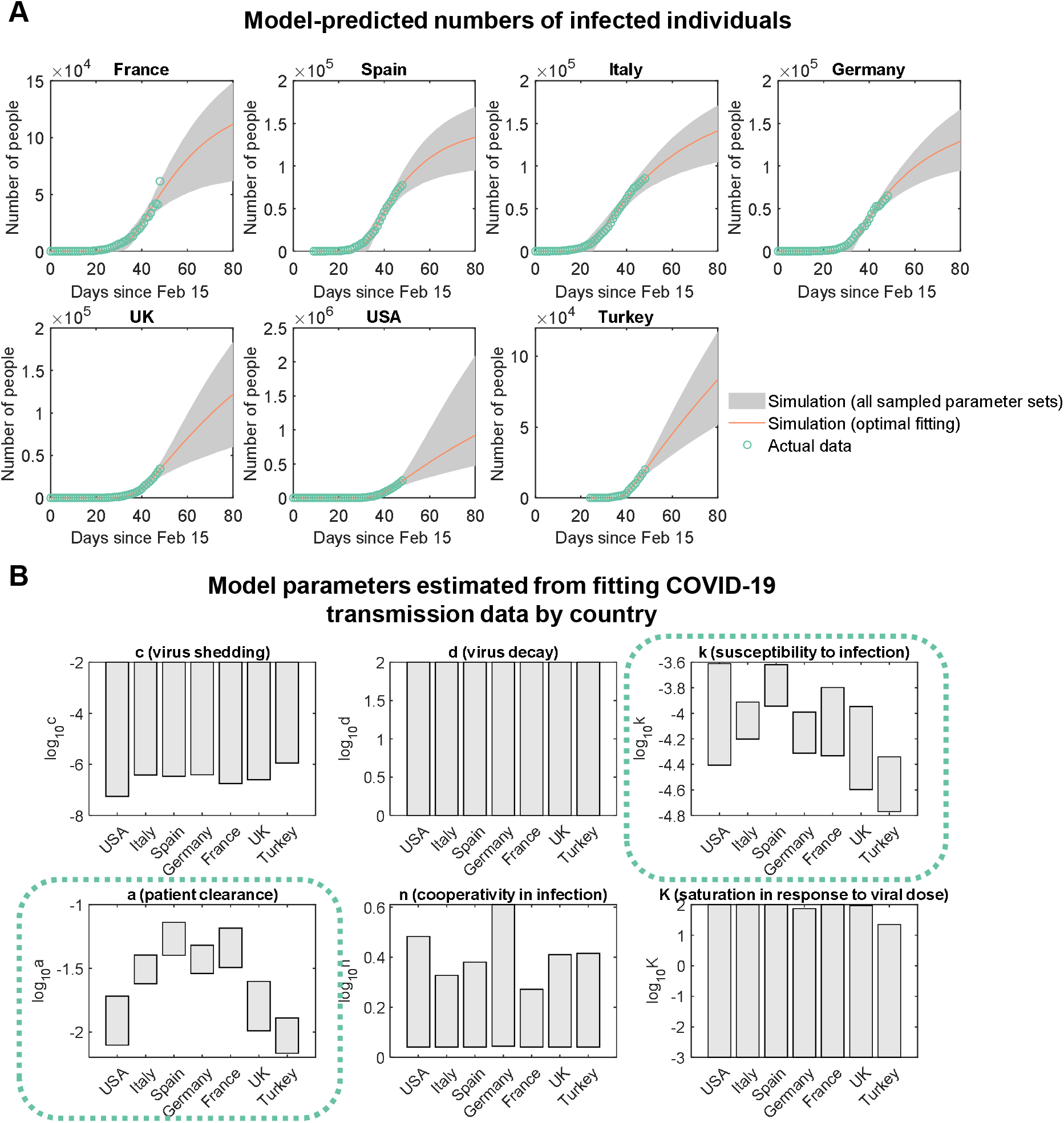
The cooperative infection model predicts country-specific transmission dynamics. A. Model-predicted numbers of infected individual during the time window of 80 days starting from February 15, 2020 and comparison between model-predictions and actual data for dates before April 3, 2020. B. Range of country-specific model parameters estimated from fitting the COVID-19 transmission data by country.

### Extremely strict interventions are needed to control the COVID-19 pandemic

We next assessed efficiency of different intervention strategies by simulating their influences on model-predicted disease transmission dynamics. We considered two scenarios of intervention: the infected individual-based intervention that targets the infected population to reduce virus shedding from the infected individuals, and the population-based intervention that limits both virus shedding from infected individuals and exposure of susceptible individuals to the virus. The two scenarios of intervention were modeled by introducing a coefficient *α* that quantifies the strength of intervention, to the cooperative infection model (Figure 3A). We considered three different levels of intervention strength ranging from strong to weak (Figure 3B), which are approximately equivalent to the approximate efficiency of N95 respirators (*α* = 0.1), surgical masks (*α* = 0.2), and homemade masks (*α* = 0.5)^28-30^. Assuming that the intervention starts at 48 days after the onset of the pandemic in the country (i.e. the date when the first COVID-19 case in the corresponding country was confirmed), we simulated the infection dynamics over a time window of 100 days starting from the onset of pandemic (Figure 3C, Supplementary Figure 2) and computed the relative number of infected individuals 100 days after the onset (Figure 3D). Notably, we found that only population-based interventions with extremely high intervention strength (*α* = 0.1, equivalent to all people wearing N95 respirators) were sufficient to effectively prevent the growth of infected population (number of infected individuals on day 100 < 15% of the number in the absence of interventions for all countries except for Turkey and 39.8% for Turkey, Figure 3D). Taken together, these results suggest that in response to the COVID-19 pandemic, extremely strict interventions that affect the entire population are indispensable to prevent the spread of the disease.

**Figure 3.**
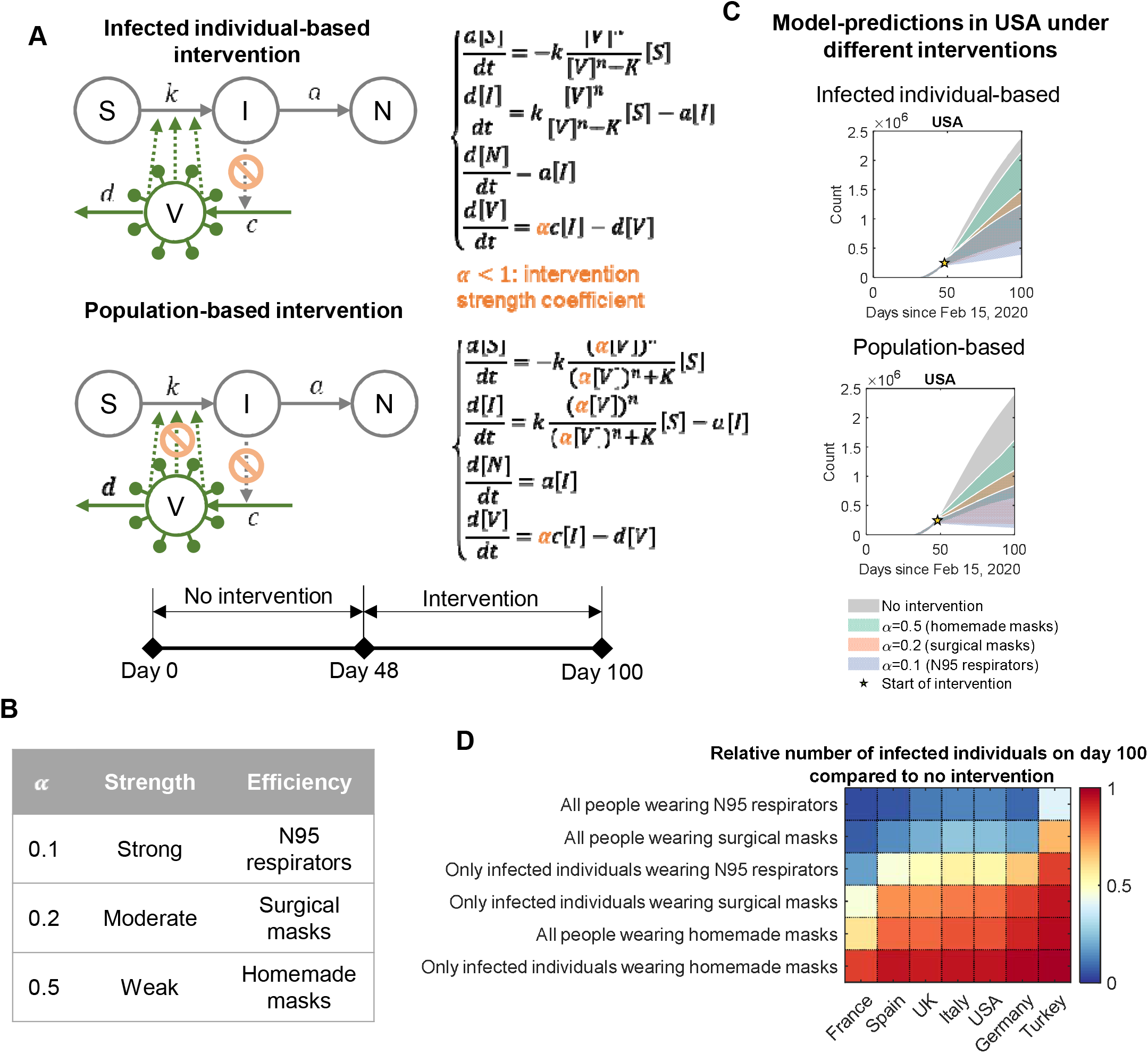
Extremely strict interventions are needed to control the COVID-19 pandemic. A. Diagram and model equations for the cooperative infection model under infected individual-based intervention (upper panel) and population-based intervention (lower panel). B. Values of the intervention strength coefficient *α* used in simulation of the interventions and their equivalent levels of virus-blocking efficiency. C. Model-predicted numbers of infected individuals before and after the intervention for both infected individual-based and population-based interventions under different intervention strengths. Shaded regions were computed from simulations based on 5,000 randomly sampled parameter sets that satisfactorily fit the country-specific transmission data. D. Relative numbers of infected individuals under interventions on day 100 since the onset of pandemic compared to the case without intervention. Values for each country were computed based on the parameter set with optimal fitting for that country.

### Cooperative infection results in resistance to public health interventions

To understand how inclusion of the cooperative infection term contributes to the response of COVID-19 transmission dynamics to interventions, we next examined how the cooperative infection term *H*, which determines the rate of infection together with the susceptibility of healthy population and number of susceptible individuals (Figure 4A), depends on the dose of viable virus. For each country, we simulated the relationship between the viral dose and the cooperative infection term using 5,000 parameter sets (Figure 4B). We found that, compared to a linear relationship, the cooperative infection term *H* has the tendency of amplifying a relatively small concentrations of virus, resulting in higher sensitivity to lower viral doses (i.e. small amount of virus is able to cause a considerable number of infections) and stronger resistance to a reduction in viral dose (i.e. moderate reduction in viral dose is insufficient to effectively prevent the infections). We further illustrate this feature of the cooperative infection model by simulating the influence of different levels of viral concentration reduction on the value of cooperative infection term (Figure 4C). We found that, under a moderate-to-strong intervention that reduces the viral concentration by 90% (the left panel), the value of the cooperative infection term was only reduced by around 40% to 70% (values ranging from 44.9% for Germany to 68.6% for UK based on parameters with optimal fitting, left panel of Figure 4C, yellow stars). This finding indicates that the majority of infections cannot be prevented by these interventions. On the other hand, under an extremely strong intervention that removes 99% of the virus (the right panel), the value of cooperative infection term was effectively reduced by the extent of more than 90% (values ranging from 93.0% for Italy and 97.3% for France, right panel of Figure 4C, yellow stars). Taken together, these results demonstrate that the cooperativity in infection, which has been found to be indispensable for the model to accurately reproduce country-specific COVID-19 transmission dynamics in our previous analysis, dampens the effects of interventions, thus requiring extremely strong interventions to effectively mitigate the pandemic.

**Figure 4.**
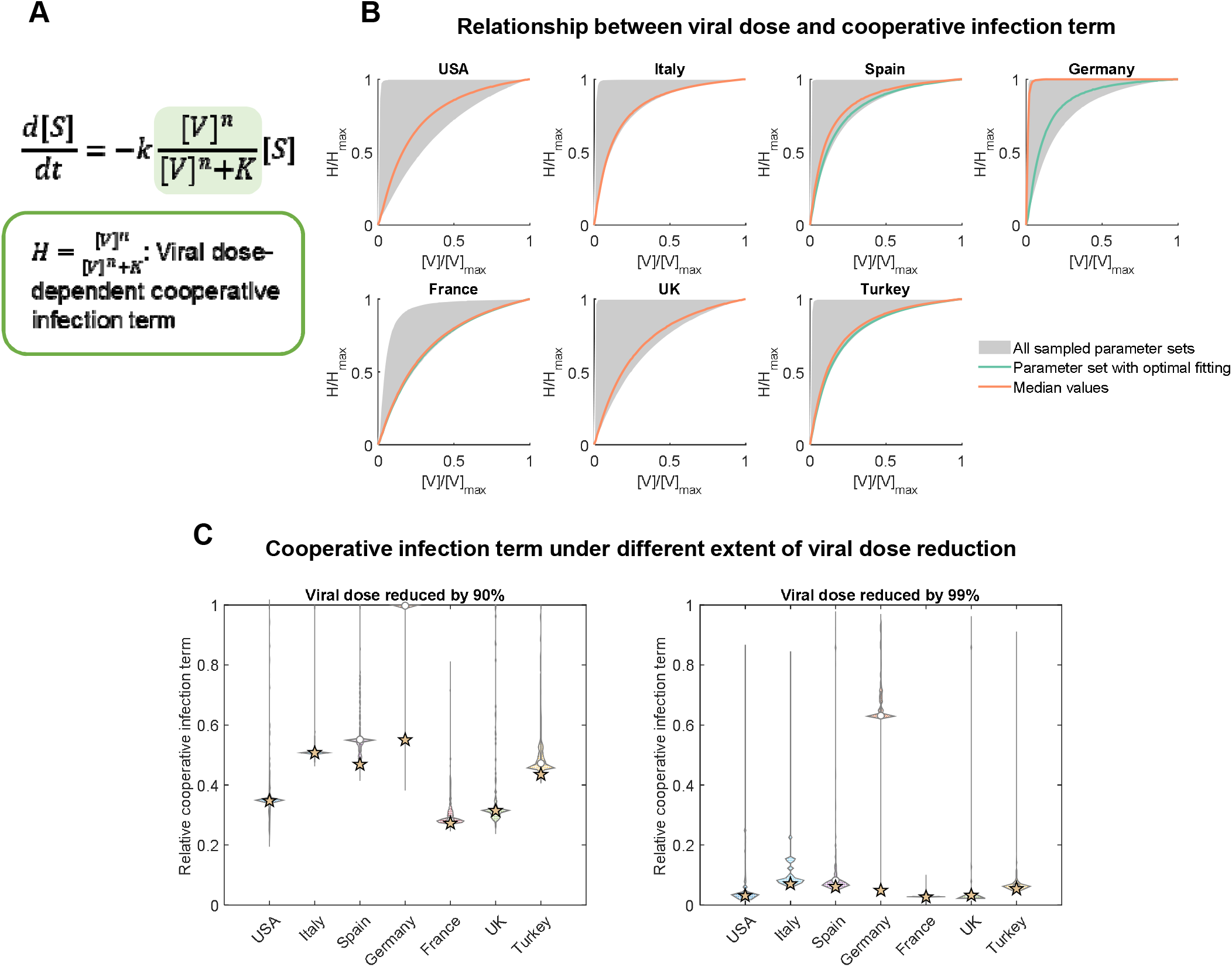
Cooperative infection results in resistance to public health interventions. A. Definition of the cooperative infection term and its relationship with the rate of infection. B. Relationship between viral dose and value of the cooperative infection term in different countries. C. Influence of reducing the viral dose by 90% (left panel) or 99% (right panel) on the value of cooperative infection term. Violin plots were drawn based on distribution of the relative cooperative infection term values computed from 5,000 randomly sampled parameter sets. Circles indicate median values based on the 5,000 simulations and stars indicate values computed based on parameter sets with optimal fitting.

### Transmission characteristics of COVID-19 in the United States of America

The USA has suffered from the highest numbers of infections and deaths among all countries, and the numbers of infections and deaths caused by COVID-19 are increasing, especially after most states have entered the initial phases of reopening in June and July of 2020. It is therefore important to understand whether the temporally varying public health responses and interventions, such as lockdowns, stay-at-home orders, travel restrictions and recommendations of wearing masks in public space, have effectively prevented the spread of COVID-19 and protected the health of the U.S. population, which reside in all sorts of neighborhoods that greatly differ with each other in cultural, social and economic factors.

To understand the state by state temporal heterogeneity in how public health interventions have shaped the COVID-19 transmission dynamics in the USA, we built state-by-state COVID-19 transmission models with the consideration that the strength of public health intervention may change with time (Figure 5A, Methods). Four different phases were modeled: an initial phase when there is no intervention; a ‘start of intervention’ phase in which the intervention strength begins to rise (i.e. the intervention coefficient *α* decreases) and finally reaches its peak value; a ‘maintenance of intervention’ phase during with the peak value of intervention strength is maintained; and, finally, a ‘lifting of intervention’ phase in which the intervention strength decreases with time. Parameters determining lengths of each phase and the rates of intervention strength changes were then estimated by fitting actual state-wide COVID-19 transmission data.

**Figure 5.**
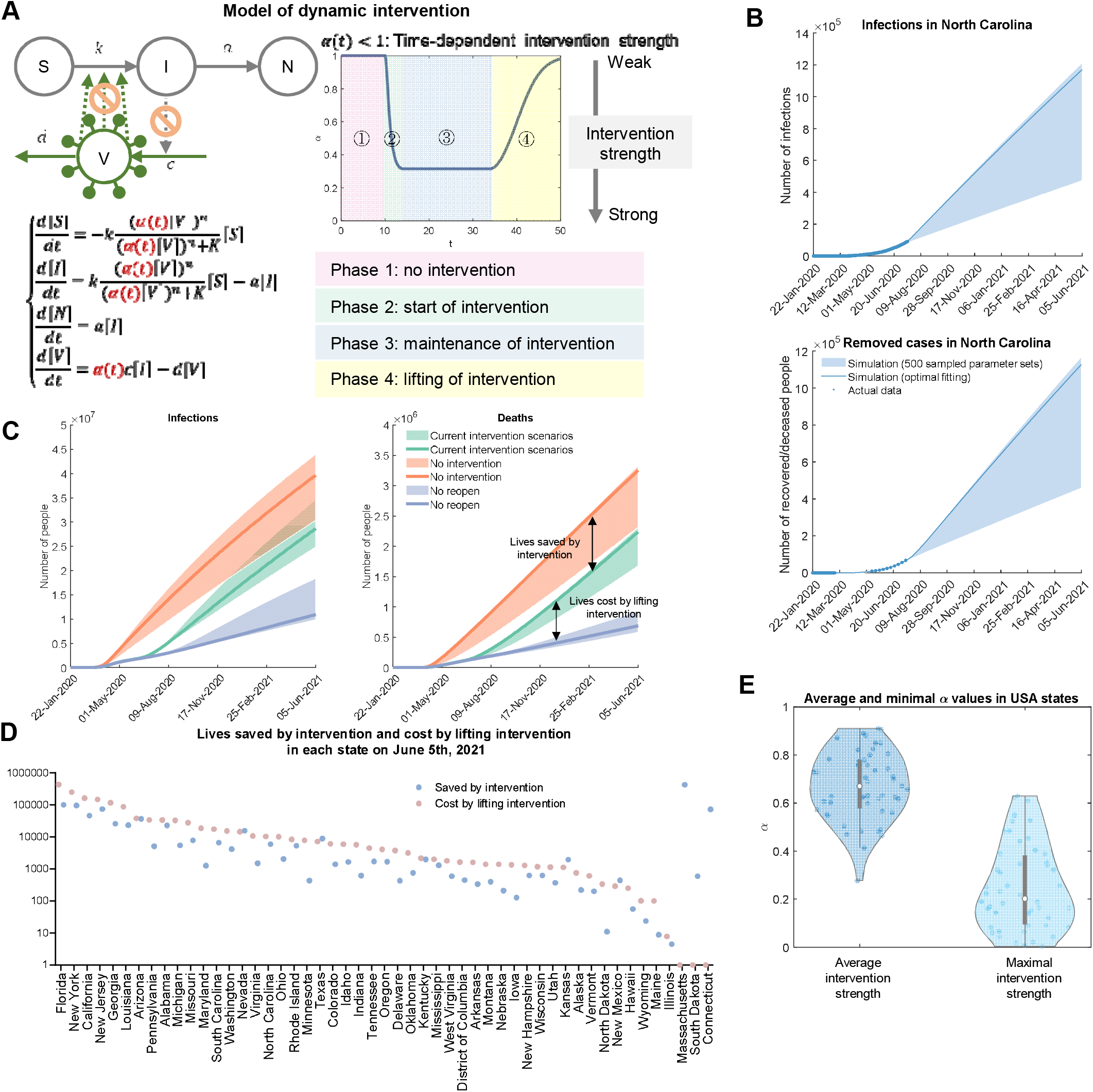
Transmission characteristics of COVID-19 in the USA. A. Diagram of the model of dynamic intervention. B. Comparison between actual and model-predicted numbers of infections and deaths in North Carolina. Shaded regions were computed from simulations based on 500 randomly sampled parameter sets that satisfactorily fit the actual transmission data. C. Model-predicted total infections and deaths in the USA under the assumption of no intervention, current intervention scenarios, or no lifting of intervention. Shaded regions were computed from simulations based on 500 randomly sampled parameter sets that satisfactorily fit the actual transmission data. D. Model-predicted numbers of lives saved by intervention and cost by lifting intervention in each territory as of June 5, 2021. Values were computed based on the parameter sets with optimal fitting for that state. E. Distributions of average and minimal values of the intervention strength coefficient *α* in territories of the USA. Values were computed based on the parameter sets with optimal fitting for that state.

We fit the model to data of COVID-19 infections, deaths and recoveries during the timespan from Jan 22, 2020 to July 15, 2020 in all 51 territories of the USA, including all 50 states and the District of Columbia (Figure 5B, S3). We found that the model satisfactorily fit the data in most of the territories (optimal cost function value < 0.005 in 40 out of 51 territories, Figure S3), confirming that the model correctly captures the spatially heterogenous COVID-19 transmission dynamics in the USA. Next, to evaluate the efficiency of public health responses in containing COVID-19 spread in each state and territory, we predicted the daily numbers of infections and deaths in the timespan from July 16, 2020 to June 5, 2021 in each state and territory, and computed the nationwide numbers of infections and deaths, assuming that the current scenarios of interventions were left unchanged (Figure 5C, ‘current intervention’), interventions were completely absent (‘no intervention’), or interventions were never lifted once the peak strengths were reached (‘no lifting of intervention’). We found that, the current interventions have saved over 500,000 lives as of July 15, 2020, and will have saved around 1 million lives at the beginning of June 2021 (5.22×10^5^ as of July 15, 2020 and 1.01×10^6^ as of June 1, 2021, Figure 5C). Notably, if in any state the interventions were never lifted, the number of lives saved by the summer of 2021 will rise to over 2.5 million (2.54×10^6^ lives saved as of June 1, 2021). In other words, the number of lives cost by lifting the interventions early can exceed the number of lives saved by imposing the interventions in the first place. This may bring into question the decision to reopen states before the pandemic is completely under control. Model-computed numbers of lives saved by intervention and cost by lifting interventions were also comparable to each other in most of the states (Figure 5D). Finally, the peak values of intervention strength coefficient *α*(*t*) in these states are distributed around 0.2 (median value = 0.202, Figure 5E, S4), while the average values of *α*(*t*) over the timespan of Jan 22, 2020 to July 15, 2020 were higher than 0.5 in almost all states (44 out of 51, Figure 5E, S4). Recalling our finding that only extremely strong interventions (i.e. *α* ≤ 0.1) are sufficient to prevent the transmission of COVID-19, these results reveal that in almost all states, the public health interventions are not strong enough to fully mitigate the COVID-19 pandemic, although they have mitigated its transmission and saved lives to some extent.

## Discussion

In this study, we have developed a mathematical model for the transmission dynamics of COVID 19 based on the concept of cooperativity in biology. The model was found to accurately reproduce the transmission dynamics of COVID-19 in seven countries that are most severely impacted by the pandemic while models without the cooperative infection were inconsistent with available data. These findings highlight the importance of cooperativity in shaping population level responses to an infectious virus and offer new insights into the transmission dynamics of infectious diseases. Notably, the cooperativity in infection leads to the resistance of virus transmission to intervention strategies that are not strong enough, resulting in increased difficulty in designing effective interventions to control the pandemic. Aggressive interventions with the strength approximately equivalent to at least requiring all individuals, no matter infected or not, to wear N95 respirators all time while in public space, are predicted to be necessary by the model. Furthermore, with the consideration of temporally varying intervention strength, we were able to extend the model to capture the temporal and spatial heterogeneity of COVID-19 transmission in the United States of America. Projection of future COVID-19 transmission dynamics and inference of intervention strengths in these states based on the models together show that the current public health interventions in the United States are not sufficient to completely mitigate the pandemic, despite being able to reduce infections and deaths caused by COVID-19 to some extent. These findings together stress the necessity of strong interventions in effectively controlling the transmission of COVID-19.

We also note that, as with any mathematical model for a complicated process, this model is not without limitations. Like all models in computational epidemiology, this model does not account for many factors that contributes to the transmission of COVID-19. First, this model assumes that there is no regional heterogeneity in the country or state of interest, and that the population is homogenously mixed, while it has been demonstrated that the susceptibility to and fatality of COVID-19 depends on many demographic factors, such as gender and age^11,31^, and life style related variables, such as smoking history^32^. Countries and state are assumed to be isolated from each other, which means no international and interstate travels occur during the time period, while imported cases likely contribute to many COVID-19 outbreaks around the world. Moreover, since a data-driven approach was used to parameterize the models, it might underestimate the severity of the pandemic because of the existence of undocumented infections due to the limitation of diagnostic capacity^33^. Other factors not included in the model, such as seasonality in transmission of coronaviruses^14^, existence of asymptotic infections^34^, and change of case definition^35^ may also affect the epidemic curve of COVID-19. Nevertheless, the model, while possibly underestimating the severity of COVID-19 due to simplification, still forecasts the rapid escalation in COVID-19 spread, and highlights the extreme urgency of stronger interventions such as contact tracing, rigorous social distancing, or the application of N95 respirators for public use by showing that moderate interventions are not likely to effectively reduce transmission due to the cooperative infection term. Modest measures such as those adopted so far in the USA unfortunately nevertheless disrupt the normal function of society and creates substantial economic and societal costs and may not be effective as planned.

In summary, this study develops a cooperative infection model that connects viral dynamics to the transmission dynamics of COVID-19 at the population level, captures the nonlinearity in the dose-response relationship in viral infection, correctly reproduces COVID-19 transmission characteristics in different countries, and, forecasts the rapid spread of COVID-19 in the absence of extremely strong interventions due to the cooperativity in SARS-CoV-2 infection. While further research and analysis are needed to improve the model and better understand the molecular mechanisms behind the cooperativity in viral infection, the predictions of this model demonstrate the indispensability of stronger interventions^14,36^ that should encourage policy makers with interests in public good.

## Methods

### Data acquisition

Time-course data for numbers of currently infected individuals (i.e. active cases) and total infected individuals (i.e. total cases, including both infected individuals and recovered or deceased individuals) in the nine countries, which were used in fitting the models, were retrieved from https://www.worldometers.info/coronavirus/. Data for total population estimates in the nine countries, which were used in estimation of the initial number of susceptible people at the onset of pandemic in each country, were obtained from the 2019 Revision of World Population Prospects (https://population.un.org/wpp/). COVID-19 transmission data in 51 territories of the USA, including daily numbers of total infected individuals, which include both currently infected individuals and recovered or deceased cases, and daily numbers of recoveries and deaths were retrieved from The COVID Tracking Project (https://covidtracking.com). Data of population in territories of the USA were obtained from The United States Census Bureau (https://www.census.gov/data/tables/time-series/demo/popest/2010s-state-total.html).

### Mathematical models for COVID-19 transmission

We develop our models based on the original SIRD model for transmission of an infectious disease:

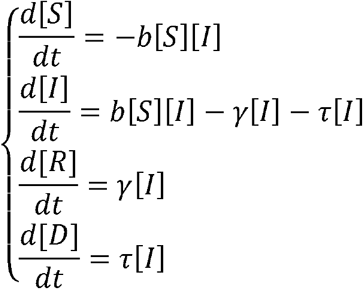

In which [S] is the total number of susceptible individuals, [I] is the total number of infected individuals, [R] is the total number of recovered individuals and [D] is the total number of deceased individuals, *b*, *γ* and *τ* are model parameters. To simplify this model, we let [*N*] = [*R*]+[*D*] denote the total number of non-susceptible (i.e. recovered or deceased) individuals. By letting *a* = *γ* + *τ*, we can then derive the equations describing the dynamics of the reduced SIRD model:

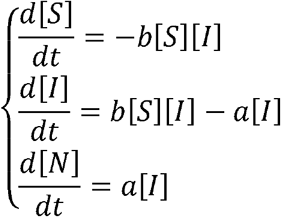

Based on the reduced SIRD model, we further include an additional term [V] quantifying the total size of viable virus pool. Dynamics of this variable depend on virus shedding from infected individuals with a rate linear to the total number of infected individuals [I], and virus decay with a rate proportional to [V]. In the absence of cooperative infection, we assume a linear relationship between the chance of a susceptible individual being infected and [V], that is, the infection of susceptible individuals happens with a rate proportional to the product of [V] and [S]. Thus, we have equations for the reduced SIRD model with virus pool:

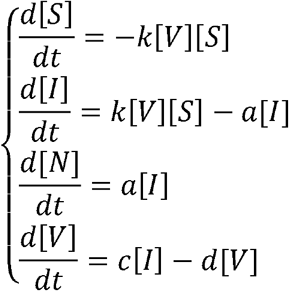

Finally, to model the cooperativity in infection of an individual, we assume that the relationship between the chance of a susceptible individual being infected and the viable virus pool size [V] follows a non-linear relationship described by a Hill function 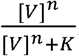. Thus, we have differential equations for the dynamics of the reduced SIRD model with virus pool and cooperative infection (i.e. the cooperative infection model):

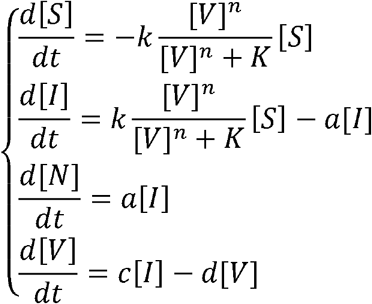

### Simulation and parameter space sampling of the models

Models were implemented with MATLAB scripts and solved using the function ode23s() for solving stiff ordinary differential equations. Model parameters for each country were estimated using differential simulated annealing, a global optimization algorithm that allows sampling of the parameter space^37^, by minimizing the cost function:

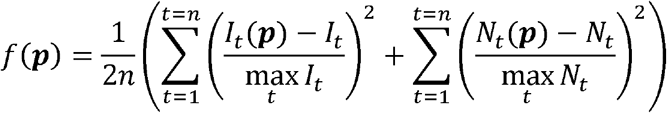

In which ***p*** is a set of parameters, *n* is the total number of time points with available data, *I_t_*(***p***) is the number of infected individuals on the *t*-th day predicted by the model with parameter set ***p***, *I_t_* is the actual number of infected individuals on the *t*-th day, *N_t_*(***p***) is the number of non-susceptible individuals on the *t*-th day predicted by the model with parameter set ***p***, *N_t_* is the actual number of non-susceptible individuals on the *t*-th day. A parameter set ***p*** is considered to generate a satisfactory fitting to the actual data if *f*(***p***) < 0.005. All parameter sets with satisfactory fitting to the data found during the sampling were kept for the following analysis. Parameters used for the differential simulated annealing algorithm were initial temperature *β*_0_ = 10^−6^, maximal temperature *β*_max_ = 10^8^, cooling rate = 1.03, Markov chain length *m* = 500.

### Sensitivity analysis

Actual numbers of infections, deaths and recoveries were assumed to be two- or five-folds of the reported values, corresponding to the assumptions that 50% or 80% of the cases were undocumented in the presence of underreporting. Hence, the numbers of infected and non-susceptible individuals on the *t*-th day used for model fitting were adjusted accordingly:

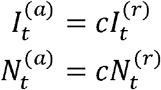

In which 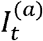 and 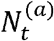denote the actual numbers of infected and non-susceptible individuals on the *t*-th day, 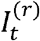 and 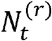 are reported numbers of infected and non-susceptible individuals on the *t*-th day. *c* is a linear factor for correction of underreporting, which has the value 2 in the case of 50% underreporting and 5 for 80% underreporting. Scripts and datasets are available in the folder “Parameter sensitivity” at the GitHub page of the project: https://github.com/LocasaleLab/COVID_19_models/tree/master/Parameter%20sensitivity.

### Simulation of interventions

Simulation of interventions was performed over a time window of 100 days starting from the onset of the first infection in each country. For each country, simulation was done with 5,000 parameter sets randomly selected from all sampled parameter sets with satisfactory fitting to the country-specific COVID-19 transmission data. An intervention was assumed to start on day 48. We first simulated the model without intervention in the time window of day 0 to day 48, and then simulated the model under the intervention from day 48 to day 100. For a parameter set ***p***, the relative number of infected individuals on day 100 under an intervention *I* was calculated as below:

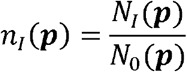

In which *N_I_* (***p***) is the number of infected individuals on day 100 with the parameter set ***p*** and under the intervention *I*, and *N*_0_(***p***) is the number of infected individuals on day 100 with this parameter set in the absence of intervention.

### Simulation of the cooperative infection term

For each parameter set, we first simulated the cooperative infection model with that parameter set in the time span from the onset of the pandemic in the corresponding country to April 3, 2020, and recorded the maximal value of viral dose [*V*], [*V*]_max_, during this time span. We then computed the values of the cooperative infection term 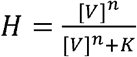 with the 101 values 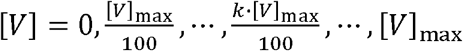. Since H is a monotonically increasing function of [V], we have the maximal value of *H* on the 101 [*V*] values: 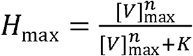. Relative values of the cooperative infection term compared to *H*_max_ were then plotted against the relative viral dose 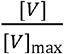 to illustrate the dependence of the cooperative infection term on viral dose.

### The model of dynamic intervention

A dynamic intervention term, *α*(*t*), was used to model the launching and loosening of public health interventions in different states of the USA:

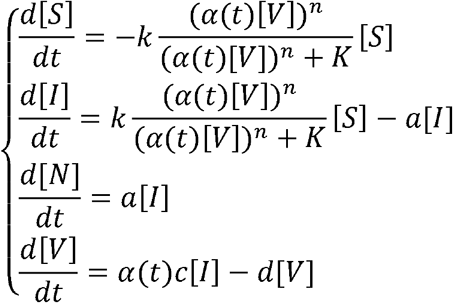

*α*(*t*) has four distinct phases depending on its quantitative relationship with the time t: first, the ‘no intervention phase’, in which *α*(*t*) has value 1, meaning that no intervention is in place during this phase; followed by the second phase, ‘start of intervention’, in which *α*(*t*) keeps decreasing until reaching its minimal value at the start of the third phase, ‘maintenance of intervention’, during which *α*(*t*) is maintained at its minimal value. The last phase is the ‘lifting of intervention’ phase in which *α*(*t*) asymptotically approaches 1 (i.e. no intervention). These dynamic behaviors of *α*(*t*) were modeled using the equation:

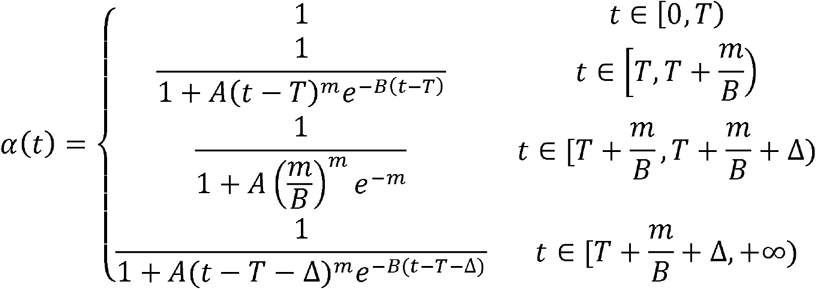

In which *A, T, m*, *B*, Δ are parameters that together determine the shape of the *α*(*t*) curve. Among these parameters, T, 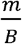 and Δ describes the lengths of the ‘no intervention’, ‘start of intervention’, and ‘maintenance of intervention’ phases. All model parameters were determined by fitting daily numbers of COVID-19 infections, deaths and recoveries in the timespan from Jan 22, 2020 to July 15, 2020 in 51 states and DC using the same fitting and sampling methods described in “**Simulation and parameter space sampling of the models**”.

### Simulation of different intervention scenarios in the USA

ODEs were solved over the time window from Jan 22, 2020 to June 5, 2021 using the function ode23s() in MATLAB R2019a. To simulate the case of no intervention, the parameter *T* (i.e. length of the ‘no intervention’ phase) was set to 10,000 days, while all other parameters were left unchanged. For the case of no lifting of intervention, the parameter Δ was set to 10,000 days, while all other parameters were unchanged. Confidence regions of the simulated curves were determined based on 500 randomly sampled parameter sets that satisfactorily fit the actual data.

### Data and code availability

Data and code used in this study are available at the GitHub page of the Locasale Lab: https://github.com/LocasaleLab/COVID_19_models. Complete results of fitting and predicting COVID-19 transmission dynamics in states of the USA are available at: https://drziweidai.com/COVID-19.html.

## Data Availability

Data and code used in this study are available at the GitHub page of the Locasale Lab. Complete results of fitting and predicting COVID-19 transmission dynamics in states of the USA are available at: https://drziweidai.com/COVID-19.html.

https://github.com/LocasaleLab/COVID_19_models

https://drziweidai.com/COVID-19.html

## Acknowledgments

JWL thanks the Marc Lustgarten Foundation, the National Institutes of Health (R01CA193256 to JWL), and the American Cancer Society 129832-RSG-16-214-01-TBE for their generous support.

---

**Figure S1.**
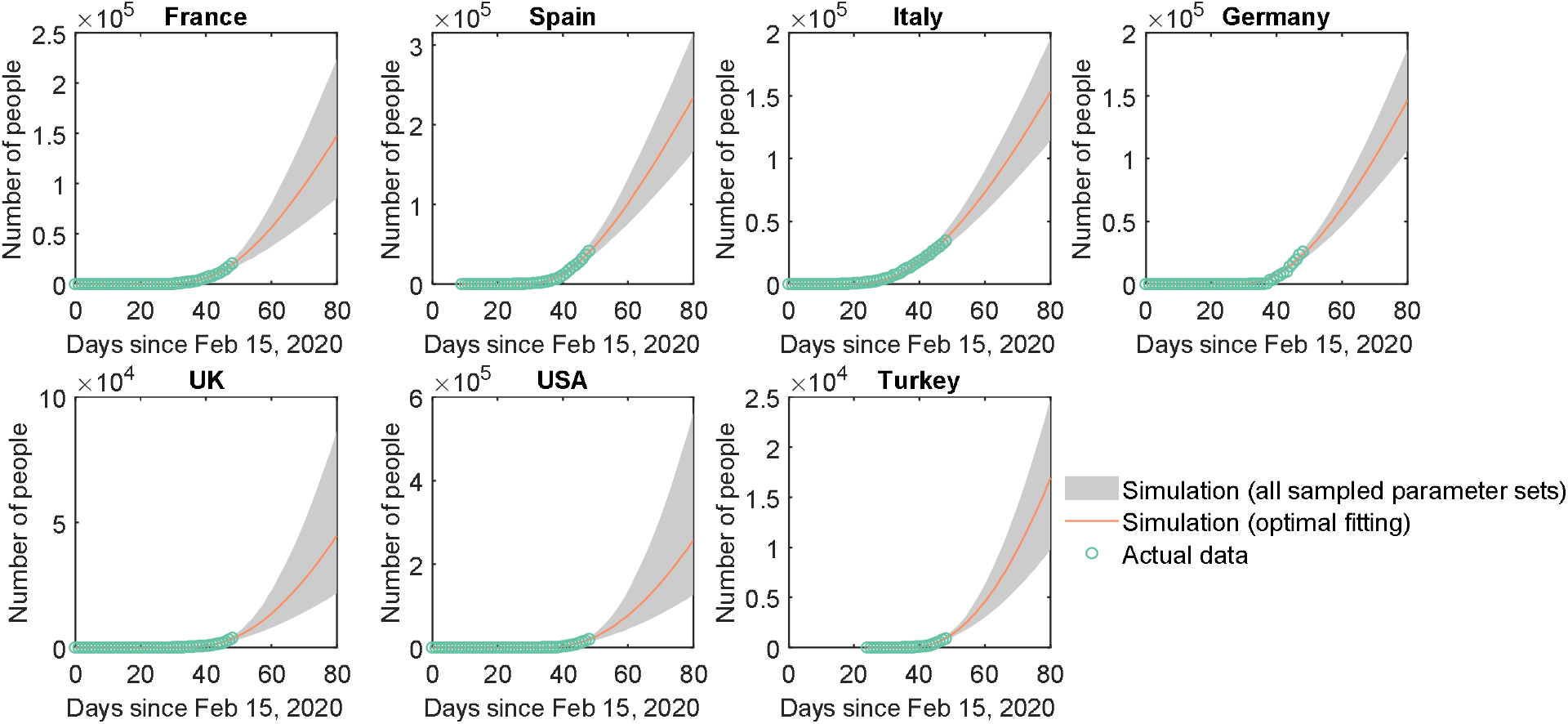
Model-predicted numbers of non-susceptible individuals.

**Figure S2.**
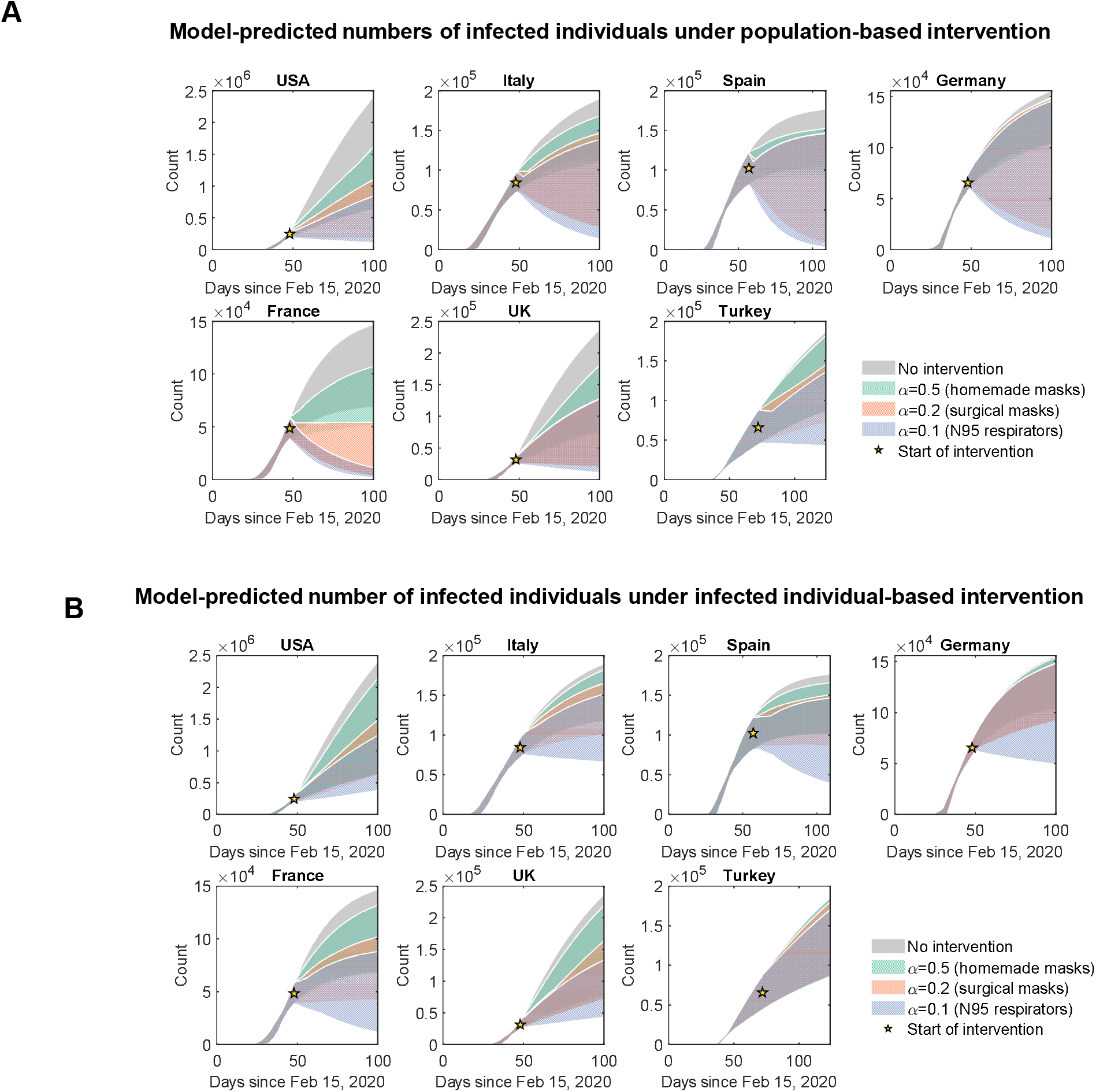
Model-predicted COVID-19 transmission dynamics under interventions.

**Figure S3.**
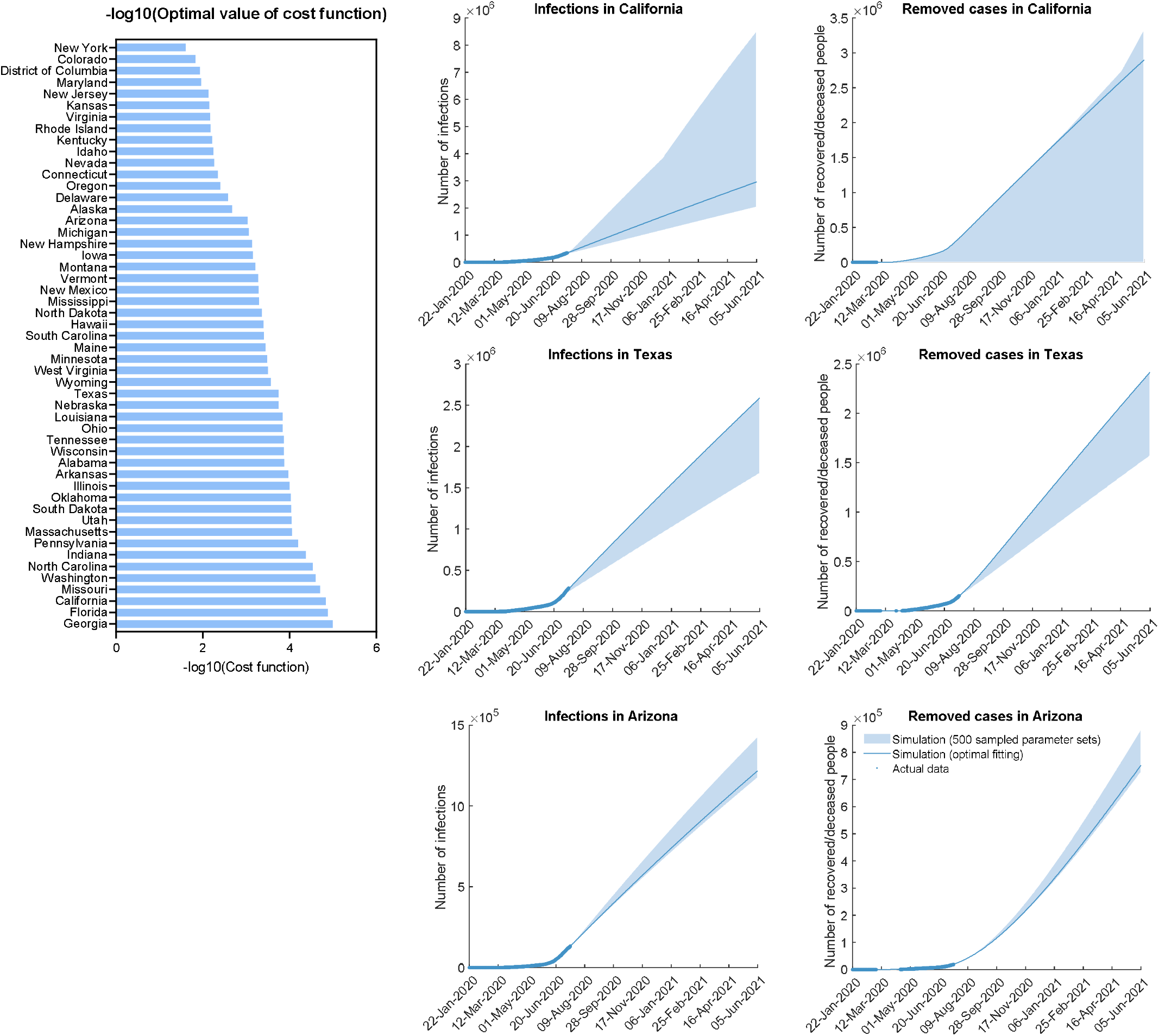
Fitting the model to COVID-19 transmission data in USA states.

**Figure S4.**
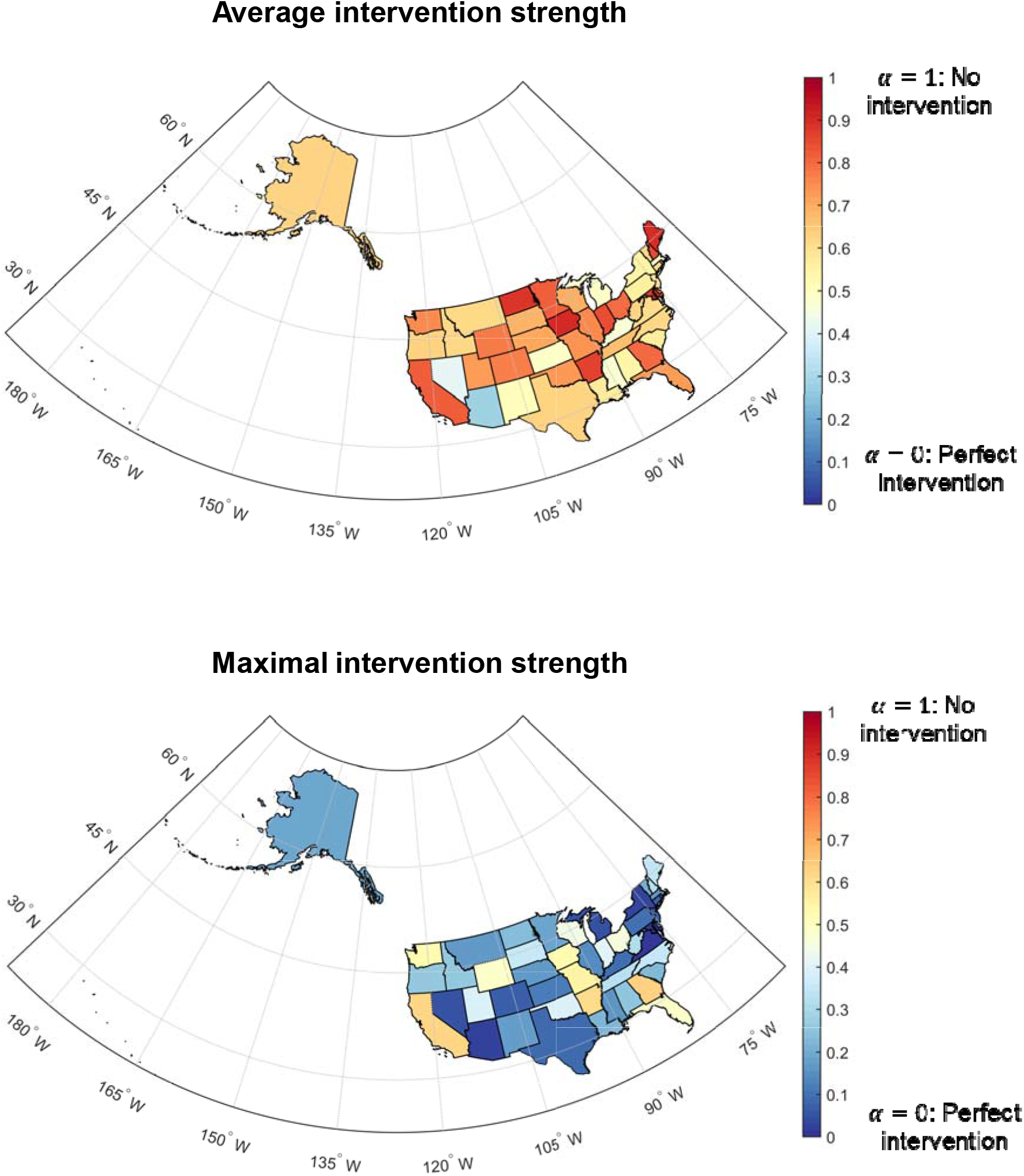
Model-inferred intervention strength in USA states.

## Notes

### Competing Interest Statement

The authors have declared no competing interest.

### Funding Statement

JWL thanks the Marc Lustgarten Foundation, the National Institutes of Health (R01CA193256) and the American Cancer Society (129832-RSG-16-214-01-TBE) for their generous support.

### Author Declarations

Not applicable for this study

### Summary of Updates

Added models for COVID-19 transmission in different states of the United States of America; revised the model to capture dynamic intervention scenarios

